# Explainable deep transfer learning model for disease risk prediction using high-dimensional genomic data

**DOI:** 10.1101/2022.01.27.22269862

**Authors:** Long Liu, Qingyu Meng, Cherry Weng, Qing Lu, Tong Wang, Yalu Wene

## Abstract

Building an accurate disease risk prediction model is an essential step in the modern quest for precision medicine. While high-dimensional genomic data provides valuable data resources for the investigations of disease risk, their huge amount of noise and complex relationships between predictors and outcomes have brought tremendous analytical challenges. Deep learning model is the state-of-the-art methods for many prediction tasks, and it is a promising framework for the analysis of genomic data. However, deep learning models generally suffer from the curse of dimensionality and the lack of biological interpretability, both of which have greatly limited their applications. In this work, we have developed a deep neural network (DNN) based prediction modeling framework. We first proposed a group-wise feature importance score for feature selection, where genes harboring genetic variants with both linear and non-linear effects are efficiently detected. We then designed an explainable transfer-learning based DNN method, which can directly incorporate information from feature selection and accurately capture complex predictive effects. The proposed DNN-framework is biologically interpretable, as it is built based on the selected predictive genes. It is also computationally efficient and can be applied to genome-wide data. Through extensive simulations and real data analyses, we have demonstrated that our proposed method can not only efficiently detect predictive features, but also accurately predict disease risk, as compared to many existing methods.

**Author summary:** Accurate disease risk prediction is an essential step towards precision medicine. Deep learning models have achieved the state-of-the-art performance for many prediction tasks. However, they generally suffer from the curse of dimensionality and lack of biological interpretability, both of which have greatly limited their applications to the prediction analysis of whole-genome sequencing data. We present here an explainable deep transfer learning model for the analysis of high-dimensional genomic data. Our proposed method can detect predictive genes that harbor genetic variants with both linear and non-linear effects via the proposed group-wise feature importance score. It can also efficiently and accurately model disease risk based on the detected predictive genes using the proposed transfer-learning based network architecture. Our proposed method is built at the gene level, and thus is much more biologically interpretable. It is also computationally efficiently and can be applied to whole-genome sequencing data that have millions of potential predictors. Through both simulation studies and the analysis of whole-genome sequencing data obtained from the Alzheimer’s Disease Neuroimaging Initiative, we have demonstrated that our method can efficiently detect predictive genes and it has better prediction performance than many existing methods.

## Introduction

Constructing an accurate disease risk prediction model is an essential step in the modern quest for precision medicine, an emerging model of health care that tailors treatments according to individuals’ profiles [1]. Over the last decades, whole genome sequencing and genome-wide association studies (GWAS) have uncovered many disease-associated genetic variants that can be used to predict genetic susceptibility. However, individual genetic polymorphisms typically explain only a small proportion of the heritability, even for traits that are highly heritable [2, 3]. It is widely accepted that most non-communicable diseases with a major public health impact are polygenic, and thus jointly modeling these genetic variants is essential for an accurate prediction model.

Polygenic risk scores (PRS) that aggregate the contributions of many single nucleotide polymorphisms (SNPs) towards the phenotype of interest has been widely used in many genetic applications, including disease risk prediction and genetic prediction of complex traits [4]. By estimating an individual’s genetic predisposition, PRS serves as a stable and measurable predictor, which can aid in the detection of diseases at an early stage and facilitate the delivery of tailored treatments. PRS, in its simplest form, is calculated as a weighted sum of SNPs that include all or more often a subset of genotyped SNPs [5–13]. The weights on SNPs can be derived from the marginal association estimated from an external GWAS data [5]. For example, [14] first used different thresholds to select SNPs from an external GWAS sample, and then generated PRS for each subject in the independent target sample. Their PRS is calculated as a weighted sum of risk alleles at selected SNPs, where the weights are the effect sizes estimated from the external sample. The weights on SNPs can also be derived from joint modeling of all SNPs, where linear mixed models and regularized regressions are commonly used [10–12]. For example, MultiBLUP predicted the phenotype by a weighted average of SNPs from multiple regions, where the weights are derived from a linear mixed model with multiple random effects to allow different genomic regions having different effect sizes [10]. More recently, the weights are proposed to be estimated using the summary statistics from GWAS [7, 9, 13]. For example, using summary statistics, lassosum estimated the weights for each SNP through solving a lasso-type of problem [7], and DBSLMM extended the Bayesian linear mixed model to handle large-scale genomic data [13].

While most PRS methods can be viewed as a weighted average of risk alleles, they differ in the assumption of distributions of genetic effects. Previous studies have shown that PRS with a flexible modeling assumption on the genetic effects can achieve more robust and accurate prediction performance across a range of phenotypes with various genetic architectures [13, 15]. While the latest developments in PRS can accommodate various types of effect size distributions [6, 13, 15], their fundamental assumption that all genetic variants act in an additive manner remains the same. However, converging evidences have shown that non-linear predictive effects (e.g., epistasis) widely exist [16]. For example, researchers have found that the protective effects of C allele within the *R1628P* variant on Alzheimer’s Diseases in Chinese Han population depend on the presence of *APOE ϵ*_4_ alleles. To capture these non-linear predictive effects, kernel functions were recently introduced into the prediction model and they have achieved various levels of successes [11, 17]. However, the performance of kernel-based methods largely depends on the pre-selected kernels, and thus can be sensitive to the underlying disease etiology. Emerging deep learning models, the state-of-the-art methods for many prediction tasks, have great potential to improve prediction models through discovering and modeling relevant features of high complexity [18, 19]. However, they generally suffer from the curse of dimensionality and provide limited insights into the genetic etiology of complex diseases, limiting their applications in the prediction of traits and disease risk. A model that can accommodate complex predictive effects and be applied to high-dimensional data is urgently needed.

While high-dimensional genomic data offers deeper insight into the genetic architecture of complex diseases, it brings tremendous challenges for the PRS construction, partially due to their huge amount of noise. Previous studies have shown that in the absence of good biological annotation, dimension reduction can be critical for an accurate risk prediction model, and indeed many existing methods only uses a subset of SNPs for PRS construction. For example, the C+T method proposed by Prive *et al*. constructed the PRS based on a subset of approximately independent SNPs obtained by clumping and p-value thresholding [9]. The recently developed DBSLMM method relies on simple regressions to identify a subset of SNPs with large-effects [13]. MKLMM and MultiBLUP split genomes into regions and selected SNPs from a subset of regions in which the effect-size variance is significantly greater than that from all other regions combined [10, 11]. MKpLMM and SARAL used penalized regression models to identify a subset of predictive SNPs based on which PRS is calculated [12, 20]. While feature selection employed in the existing PRS methods reduces the impact of noise, the pre-selected features may only have sub-optimal prediction performance, as the objectives for feature selection and prediction modeling are not the same. For example, C+T procedure uses p-value thresholding to select SNPs [9]. However, p-value is a function of effect size and sample size, but neither of them has one-to-one correspondence with prediction accuracy. Therefore, important predictors can be missed due to the pre-selection. In addition, the existing feature selection methods are not efficient in extracting and modeling SNPs of high complexity (e.g., interaction effects).

While deep learning models are natural choices for capturing non-linear predictive effects, most of them are not designed for the dimension reduction purpose. Feature importance scores, such as those used in knockoff and Gaussian mirror models [21–28], have great potential to be adapted for detecting predictive features. However, both knockoff and Gaussian mirror models double the dimension of input due to the construction of additional variables, and thus are not directly applicable for analyzing high-dimensional data. Permutation-based feature importance score methods require model re-fit, and they tend to be computationally expensive, especially for complex models (e.g., deep networks). Gene is a functional unit of DNA, and thus selecting predictive features at the gene level not only reduces the computational complexity, but also improves the model interpret-ability. However, existing feature importance scores designed for deep learning models mostly focus on individual features [23, 26, 27], and thus not directly applicable for identifying predictive genes. Group knockoff models are among one of the few methods that have the potential for identifying predictors at the gene level [29]. However, like knockoff models, the performance of group knockoff models can be sensitive to the misspecification of the conditional distribution of features to be tested and the dimension of the input layer has doubled due to the construction of knockoffs.

To address these limitations, we developed a deep transfer learning model for the calculation of PRS. We first developed explainable group-wise feature importance scores for the dimension reduction, where genes that harbor features with various types of predictive effects can be efficiently identified with controlled false positive rate. We then used the idea of transfer learning to build an explainable deep transfer learning model for the PRS construction, where information from feature screening is directly incorporated. In the following sections, we first presented our proposed screening rule and the deep transfer learning model, and then examined the selection and prediction performance of our method. Finally, we applied our method to predict AV45 and FDG using data obtained from the Alzheimer’s Disease Neuroimaging Initiative [30].

## Materials and methods

Set-based analyses that aggregate signals from all features within a set, have greatly facilitated the detection of disease-associated regions. Using a similar idea in [29], we first developed a group-wise feature importance score to screen the genome to select predictive regions (e.g., gene), and then used the idea of transfer leaning to build a predictive model, where information from feature screening is directly incorporated.

### Feature selection

For *n* i.i.d. sample, let ***Y*** = (*Y*_1_,…, *Y*_*n*_) be the phenotype and ***X***_*k*_ = (***X***_1*k*_,…, ***X***_*nk*_) be the genotype for the *k*th genomic region (i.e., gene), where 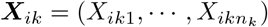 with *n*_*k*_ being the number of genetic variants for the region. *Y*_*i*_ can be either a real number in a regression setting or an index from *{*1, 2,…, *C}* in a classification problem. We further split the data into two subsets ℐ_1_ and ℐ_2_, and let 𝒟_*k*_ = *{*(***X***_*ik*_, *Y*_*i*_) : *i* ∈ *ℐ*_*k*_*}, k* = 1, 2.

The goal for feature screening is to identify regions (e.g., gene and pathways) that harbor predictive genetic variants, where the effects can be linear and/or non-linear. As deep neural network (DNN) is the state-of-the-art method for modeling features with non-linear effects, we propose to first fit a simple DNN model (e.g., multi-layer perceptron) for each region, and then construct a group-wise feature importance score to gauge the predictive importance of each region. Let *f*_*k*_(***X***_*k*_) be a predictive model built based on region *k* trained on 𝒟_1_. The model *f*_*k*_(***X***_*k*_) can be viewed as a conditional mean (i.e., *E*[*Y* |***X***_*k*_]) for continuous traits, or a conditional probability (i.e., *P* (*Y* |***X***_*k*_)) for categorical outcomes. Let *L*(*Y*_*i*_, *f*_*k*_(***X***_*ik*_)) denote a chosen loss function. For example, mean square error and cross entropy can be used as loss functions for continuous and binary outcomes, respectively. We propose to construct the group-wise feature importance score, denoted as Δ_*k*_, to gauge the predictive importance of region *k*:

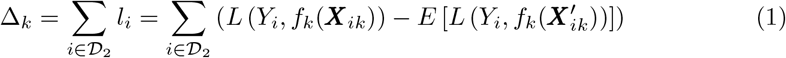

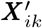 represents the permuted data that is obtained via randomly shuffling the labels, and thus the permuted genetic data can maintain their intrinsic structures (e.g., linkage disequilibrium).

By definition, 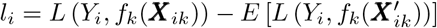. Therefore, the proposed group-wise feature importance score is obtained by comparing the loss derived from the observed and permuted data that is known to be not predictive. Intuitively, if region *k* is not predictive, then the difference in loss between observed and permuted data should be around zero, and thus Δ_*k*_ is expected to be 0. Alternatively, if region *k* is predictive, then the loss in observed data is expected to be smaller than that from permuted data, leading to Δ_*k*_ *<* 0. Therefore, Δ_*k*_ *<* 0 measures the predictive power of region *k*, and a smaller negative value indicates higher predictive power.

We propose to test whether Δ_*k*_ is significantly smaller than 0 to determine whether region *k* is predictive, and used the significance as a proxy to gauge the predictive power of region *k* for a given DNN model. Therefore, we perform a one-sided hypothesis test:

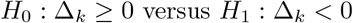

Under the null, we assume that *l*_*i*_ (*i* ∈ *𝒟*_2_) comes from the same distribution and var(*l*_*i*_) *<* ∞, as |*𝒟*_2_| → ∞. Therefore, we have 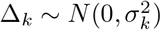, where |*𝒟*_2_| denotes the cardinality of 𝒟_2_ and 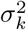 is estimated empirically.

Although DNNs that can capture both linear and non-linear effects have good generalization properties, they tend to be over-parameterized and can be overfit on the training data (i.e., 𝒟_1_). Therefore, the differences in loss are proposed to be evaluated on the validation data (i.e., 𝒟_2_). However, randomly splitting the data into two subsets (i.e., 𝒟_1_ and 𝒟_2_) once can create chance finding, and thus we use the idea of *K*-fold cross-validation to define the overall test statistics as

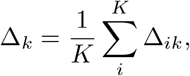

where Δ_*ik*_ is calculated using equation 1 based on the data from the *i*th cross-validation. Given each Δ_*ik*_ asymptotically follows a normal distribution (i.e 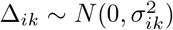), we have 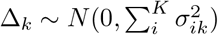 and thus the predictive significance of region *k* can be evaluated accordingly.

There are two fundamental differences between our proposed feature selection method and other screening rules: 1) By applying the fitted model *f*_*k*_(·) on the permuted data 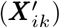, we eliminate the need to refit a new model on each permuted data, making it easy to evaluate the loss difference. The main advantage of such a strategy is its computational efficiency, which is of great importance for complex DNN models. Indeed, refitting hundreds of DNNs for each region is computationally prohibited. 2) Our proposed screening method aligns well with the downstream prediction task. Unlike many existing methods that treat variable screening (e.g., SIS [31] and HSIC-Lasso [32]) and predictive modeling as independent processes, the proposed group-wise feature importance score is designed to measure the predictive power for each region, and thus is consistent with the goal of prediction modeling. The p-values derived from our proposed test can be viewed as relevant importance for the prediction task, and thus can provide practical guidance for feature selection. In addition, the proposed method allows for considering the joint predictive effects from all features within the region, which not only makes it possible to capture features with complex effects (e.g., interaction), but also greatly facilitates model interpretation.

### Prediction modeling

Deep learning models are the state-of-the-art method for capturing and modeling features of high complexity. However, they generally suffer from the curse of dimensionality and lack of interpretability. While our proposed feature selection can reduce data dimension substantially, training a DNN with pre-selected features can still be computationally expensive and need a huge amount of memory. In addition, DNNs may fail to capture the infinitesimal effects that have been often assumed by existing PRS methods [33]. In this work, we propose to design a new network architecture, where DNNs built from feature screening are combined based on the idea of transfer learning and a background node is further added to capture the infinitesimal effects.

An illustrative figure of our proposed idea is shown in Fig 1. The background node (i.e., 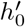) is designed to capture the infinitesimal effects and is obtained by fitting a gBLUP model [33]. As the objective function in feature screening is the same as that in the final prediction task, the networks trained during the feature screening process are informative for the final prediction task. Therefore, we use the idea of transfer learning to build the final prediction model. We propose to treat the last hidden layers of pre-trained models obtained from feature screening as the input, and stack the newly added hidden layers on top to model the joint effects from these selected genes.

**Fig 1.**
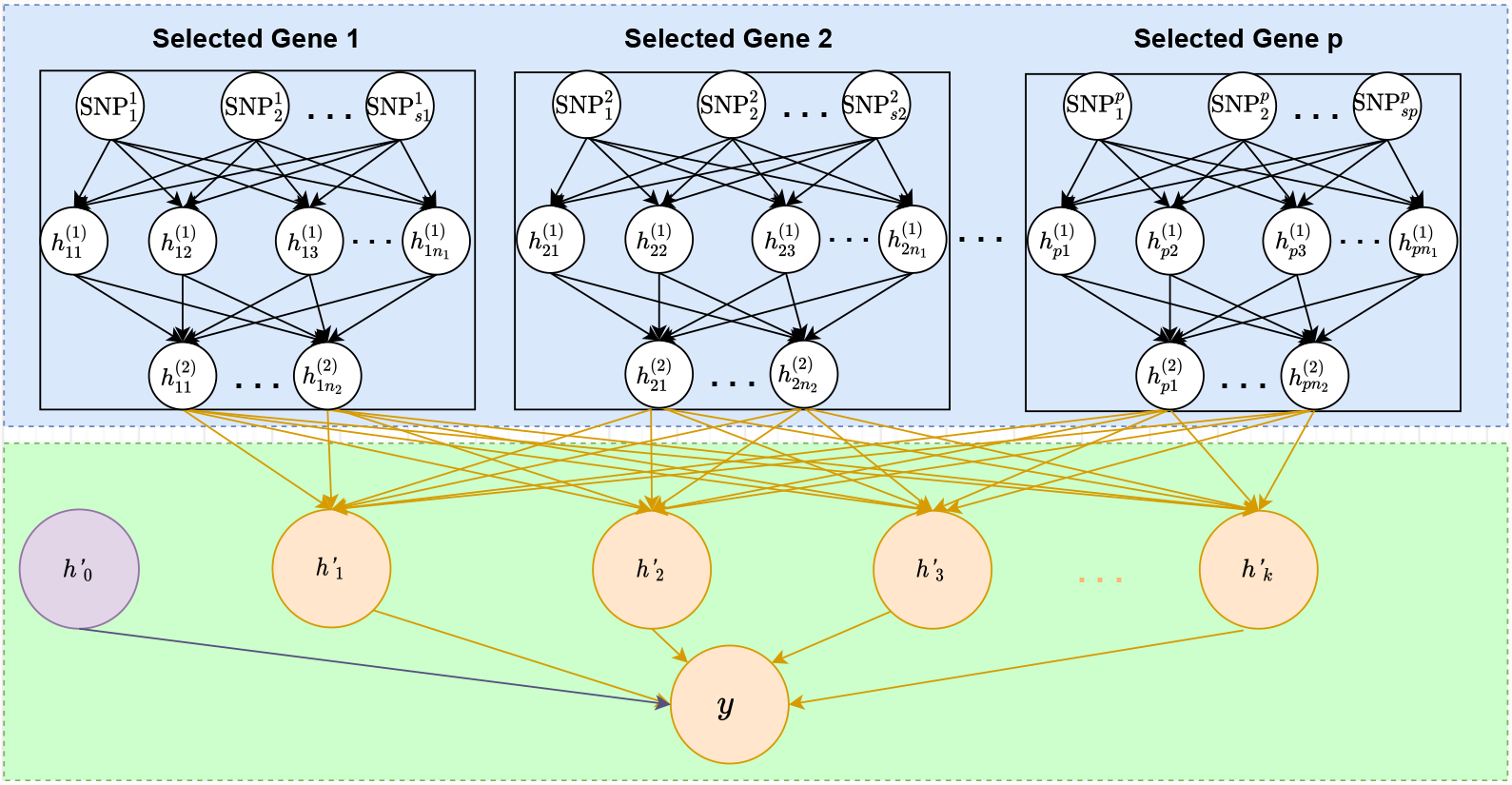
An illustrative figure of the architecture of the proposed transfer-learning-based deep network. The blue box: DNN models obtained from feature screening and the corresponding parameters are fixed. **The green box**: the background node 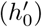 capturing the infinitesimal effects and the newly added hidden layers designed to model the joint effects from selected genes. The parameters associated with the background node and the newly added hidden layers are estimated.

Specifically, let *p* denote the number of genes selected based on our proposed group-wise feature importance score presented in the previous section, and 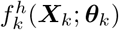 represents the last hidden layer of the DNN model trained on gene *k*, where ***θ***_*k*_ is a vector of the associated model parameters. Our proposed prediction model can be presented as

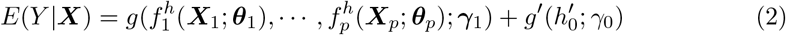

where *g*(; ***γ***_1_) is a function associated with the added hidden layers with parameters ***γ***_1_, and *g*′(; *γ*_0_) is a function for the background with parameter *γ*_0_. Same as transfer learning, we keep the parameters associated with pre-trained models unchanged (i.e., ***θ*** is fixed in the network), and only estimate parameters for the background (*γ*_0_) and newly added hidden layers (i.e., ***γ***_1_). Therefore, the proposed transfer learning model substantially reduced the number of parameters as compared to a DNN with the same architecture, where (*γ*_0_, ***γ***_1_, ***θ***_1_,, ***θ***_*p*_)′ needs to be estimated. Similar to the feature screening process, standard loss function (e.g., mean square error and cross entropy) and optimization technique (e.g., Adam) are used for parameter estimation.

The algorithm of our proposed framework, including both feature screening and prediction modeling, is depicted in algorithm 1. There are five major advantages of our proposed framework: 1) It streamlines the dimension reduction and prediction modeling processes. By making the objective functions for feature screening and prediction modeling the same, our method reduces the chance of overlooking important predictors, and thus has great potential to improve prediction accuracy. 2) Both feature screening and prediction modeling are built based on DNN models, and thus it has the natural advantages of capturing and modeling features of high complexity, making it easy to account for both linear and non-linear predictive effects. 3) Our method selects predictors at the gene level, and thus has better interpretability as compared to many existing DNNs that are completely black-box nature. 4) By using the idea of transfer learning, we have substantially reduced the number of parameters in our prediction model, and thus improve the memory and computational efficiency. 5) The proposed framework is very flexible, and can easily accommodate various model assumptions. For example, if interplay among genes is considered, the network shown in Fig 1 can be used. On contrary, if only interactions within genes are considered, the network in Fig 2 can be used instead, where fully connected layer is replaced by a pre-specified structure. In addition, unlike existing PRS that either assumes diseases are caused by infinitesimal effects or large isolated effects, our model can consider both conditions simultaneously. For example, when ***γ***_1_ is estimated to be zero, our model is equivalent to gBLUP that assumes the infinitesimal effects [33]. Similarly, when 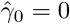, our model is equivalent to a sparsity regression model, which assumes diseases are affected by a few genes with large effects [34]. When both *γ*_0_ and *γ*_1_ are not zeros, our method models both the effects from isolated large predictors as well as the infinitesimal effects.

**Fig 2.**
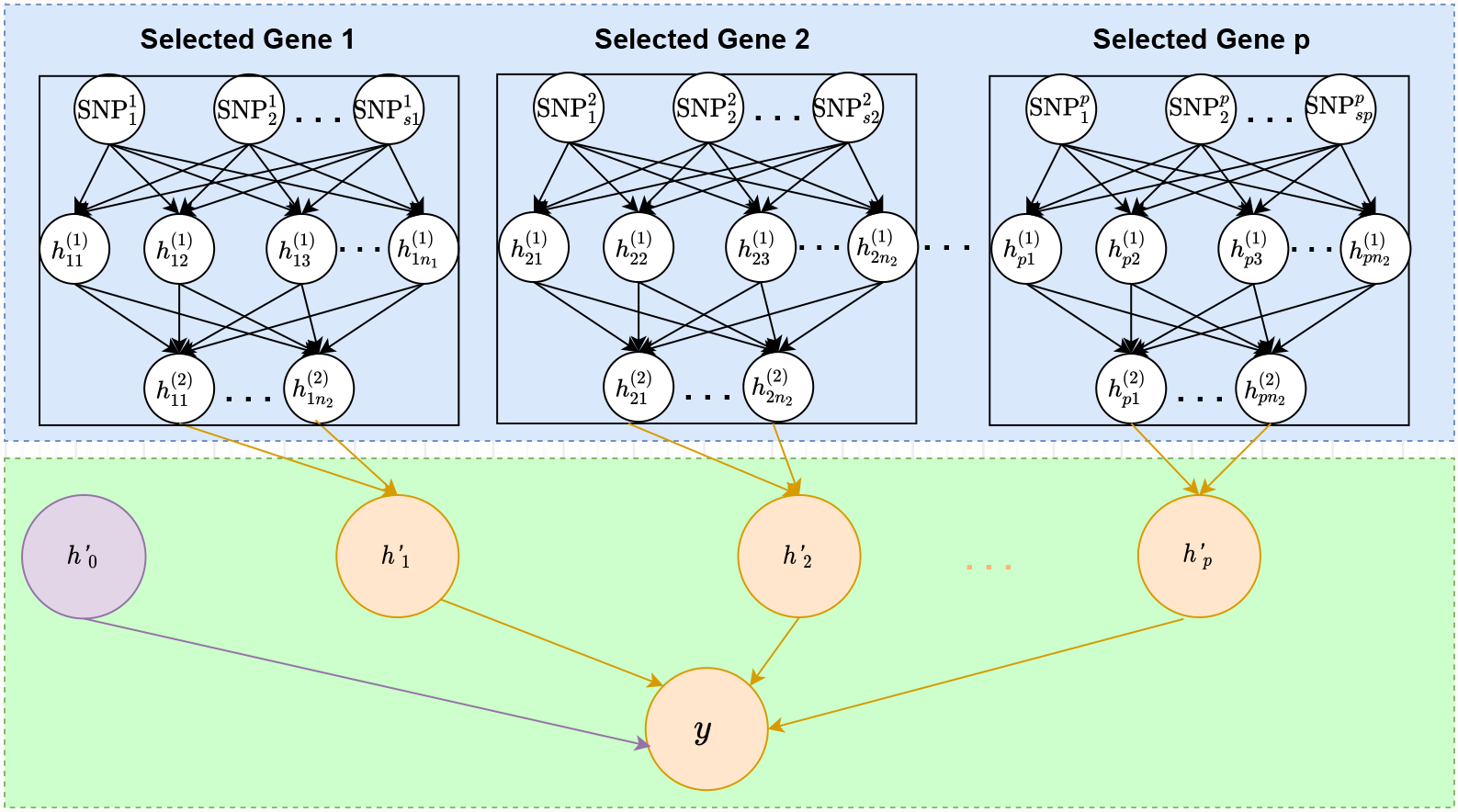
An illustrative figure of the architecture of the proposed transfer-learning-based deep network, where no interaction between genes is assumed. The blue box: DNN models obtained from feature screening and the corresponding parameters are fixed. **The green box**: the newly added hidden layers, a background node, and their associated parameters that need to be estimated.

#### Algorithm 1 Deep Neural Network-based Prediction Model

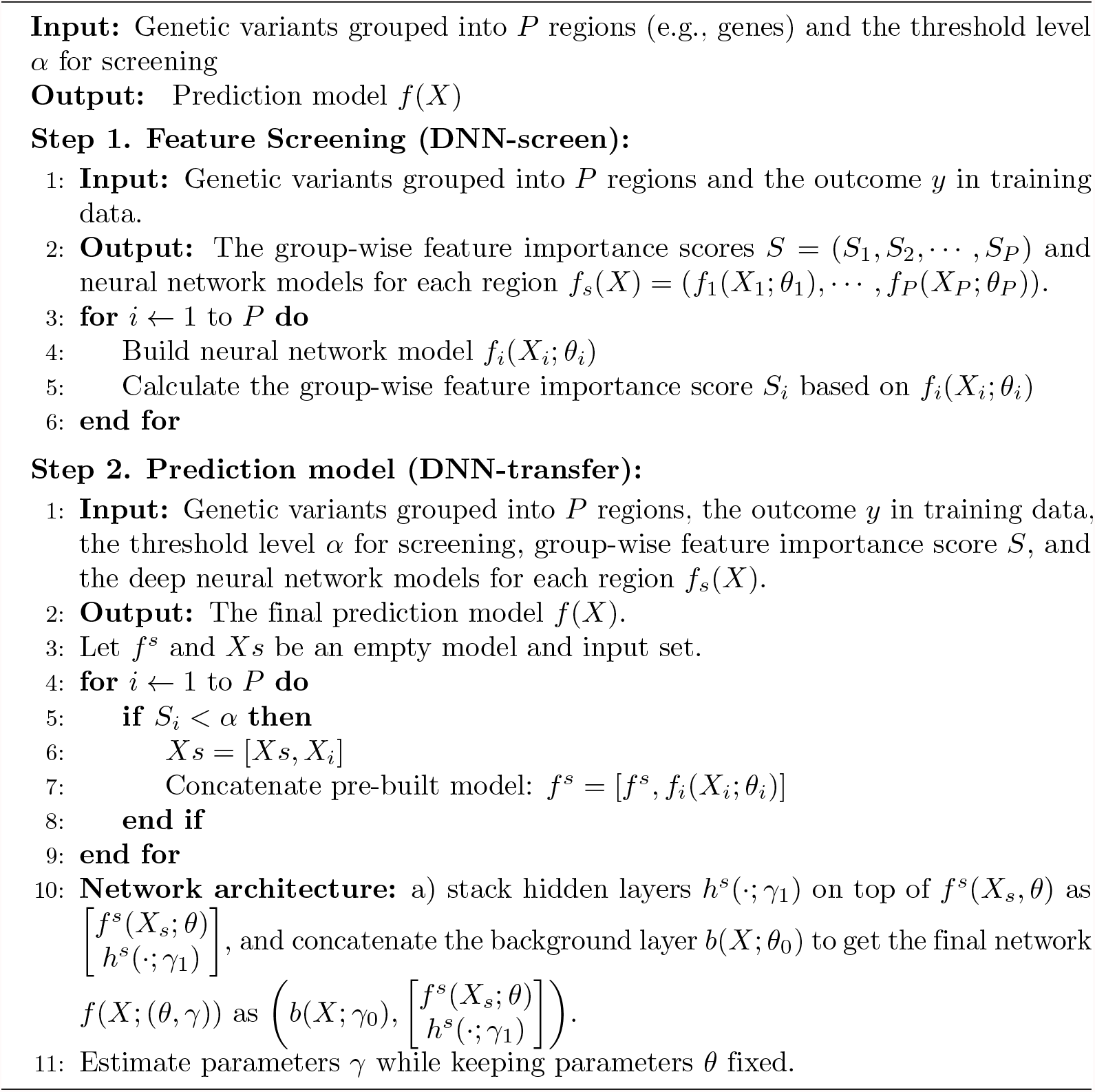

### Simulations

We conducted a set of simulations to evaluate the performance of our method, including feature selection and prediction modeling. To mimic the human genome, genotypes were directly drawn from the UK Biobank data, where unrelated white British individuals with missing genotype rate less than 5% were included. We cut the genome into genes based on GRCh37 assembly, and excluded variants that meet any of the follow criteria: 1) minor allele frequencies *<* 1%; 2) INFO score *<* 0.8; and 3) missing rate *>* 5%.

### The evaluation of feature selection

We evaluated the performance of feature selection based on power and type I error. We considered a total of ten genes with six harboring only noise variants, and simulated the mean using the remaining four genes as:

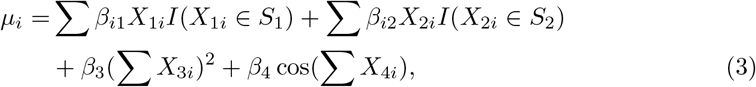

where 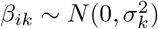 and *S*_*k*_ indicates the sets of causal variants for gene *k* (*k* ∈{1, 2 }). We set 90% and 10% of genetic variants in the first and second genes to be causal (i.e., *I*(*X*_1*i*_ ∈ *S*_1_) = 90% and *I*(*X*_2*i*_ ∈ *S*_2_) = 10%). As shown in equation 3, in addition to genes with only additive effects, another two genes that harbor non-linear effects, including pairwise interactions and a cosine function, were considered. We simulated both normally distributed and binary outcomes as:

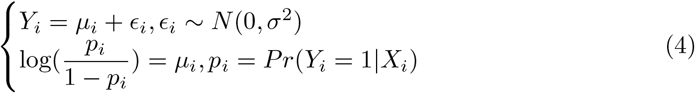

We considered sample sizes of 1,000 and 10,000 in our simulations, and the corresponding effect sizes are shown in S1 Table. For each setting, we did 5000 Monte Carlo simulations. For our proposed feature selection (denoted as DNN-screen), DNN is set as a multi-layer perceptron that has 2 hidden layers with 50 and 10 nodes for the first and second layers, respectively. To control over-fitting, we also added a dropout layer after the first hidden layer, where the dropout rate was set within the recommended range of 0.2 to 0.8. A total of 100 epoch was used for each model training. For comparison purposes, we analyzed the same data using a widely-used set-based method, including SKAT with a linear kernel (denoted as SKAT-linear) and SKAT-optimal that optimally combines the burden test and SKAT [35, 36]. We also analyzed each simulated data by first using a single variant test, and then derived the region-based p-value using the ACAT, a recently proposed Cauchy-statistics-based method [37].

The type I errors for both continuous and binary outcomes are shown in Table 1. For all methods considered, the type I errors are well controlled under the significance level of 5% and 1%. The power under the significance levels of 5% and 1% is shown in Fig 3 and S1 Fig, respectively. Not surprisingly, our proposed DNN-based group-wise feature importance score significantly outperformed the other three methods when the predictive effects are non-linear. This is mainly because neural networks are particularly powerful at capturing non-linear effects without the need of pre-specifying relationships between predictors and outcomes. When causal genetic variants act in a linear additive manner, our proposed DNN-screen performs similarly to ACAT that combines p-values from all variants within the region [37]. While SKAT with a linear kernel has similar performance to the proposed method when most of the variants are causal, its power tends to reduce when only a small proportion of the variants are associated. Although ACAT method that is mainly driven by small p-values tends to perform better than SKAT-based methods under all situations considered, it performed worse than our method when the predictive effects are non-linear. Nevertheless, as shown in Fig 3 and S1 Fig, our proposed DNN-based group-wise feature importance score performs similarly to the existing widely used set-based methods when the predictive effects are linear, and it significantly outperformed these methods when non-linear effects present.

**Table 1.** The comparisons of type I errors based on 5000 Monte Carlo simulations

|  | No | Continuous |  |  |  | Binary |  |  |  |
| --- | --- | --- | --- | --- | --- | --- | --- | --- | --- |
|  |  | DNN-screen | SKAT-linear | SKAT-optimal | ACAT | DNN-screen | SKAT-linear | SKAT-optimal | ACAT |
| $\alpha = 0.05$ | 1000 | 0.049 | 0.050 | 0.052 | 0.059 | 0.049 | 0.046 | 0.049 | 0.041 |
|  | 10000 | 0.054 | 0.053 | 0.056 | 0.055 | 0.048 | 0.049 | 0.050 | 0.050 |
| $\alpha = 0.01$ | 1000 | 0.009 | 0.012 | 0.010 | 0.016 | 0.011 | 0.009 | 0.010 | 0.007 |
|  | 10000 | 0.010 | 0.010 | 0.012 | 0.013 | 0.012 | 0.012 | 0.011 | 0.012 |

**Fig 3.**
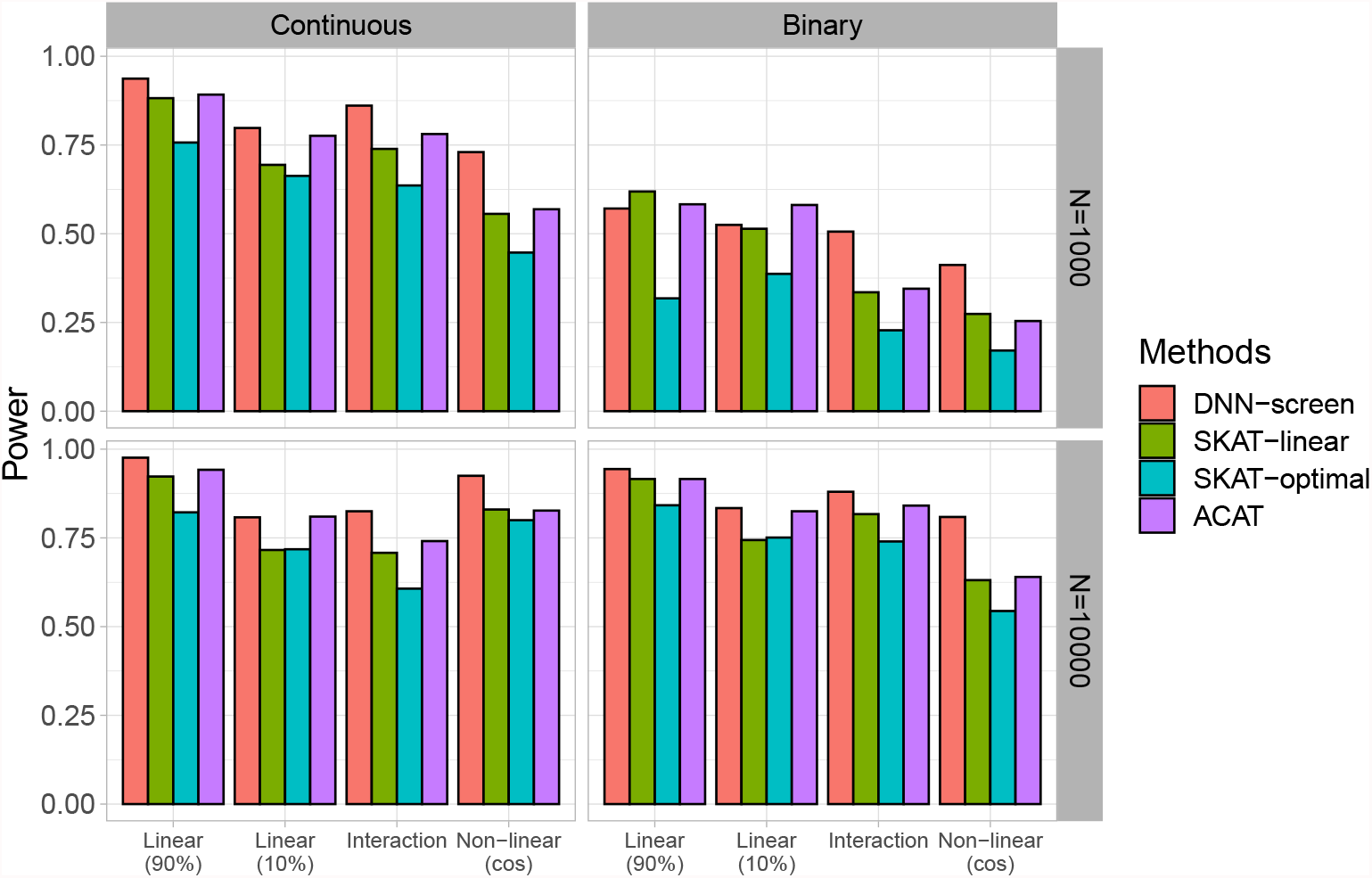
The comparisons of power under 5% significance level based on 5000 Monte Carlo simulations. Linear (90%): 90% of genetic variants on the causal gene is predictive. Linear (10%): 10% of genetic variants on the causal gene is predictive. Interaction: pairwise interaction effects. Non-linear (cos): genetic variants on the causal gene affect the outcome through a cosine function.

### The evaluation of prediction modeling

To evaluate the prediction performance, we also simulated both binary and continuous outcomes using equation 4, where the conditional mean (*μ*_*i*_) included both linear and non-linear predictive effects:

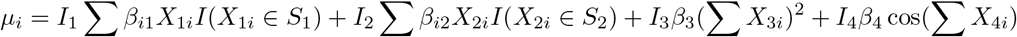

We considered three types of disease models, including 1) *S*_1_: linear effects only model (i.e., *I*_1_ = *I*_2_ = 1 and *I*_3_ = *I*_4_ = 0), 2) *S*_2_: non-linear effects only model (i.e., *I*_1_ = *I*_2_ = 0 and *I*_3_ = *I*_4_ = 1), and 3) *S*_3_: both linear and non-linear effects (i.e., *I*_*i*_ = 1, ∀*i*). Similar to simulation 1, we set *I*(*X*_1*i*_ ∈ *S*_1_) = 90% and *I*(*X*_2*i*_ ∈ *S*_2_) = 10%. The details of effect sizes are summarized in S2 Table. For each model setting, we gradually increased the number of noise genes from 6 to 96 (i.e., the total number of genes increases from 10 to 100), and did 1000 Monte Carlo simulations. To evaluate the prediction accuracy, Pearson correlations and the area under the curve (AUC) are used for continuous and binary outcomes, respectively. For our proposed framework, we first used feature selection to screen predictive genes. We then selected those that met the p-value threshold, and treated them as the input for prediction modeling. The network architecture of our model is the same as Fig 1, where the DNNs in the screening phase were treated as pre-trained models and two fully connected hidden layers were stacked on top. The number of hidden nodes for the first and second hidden layers were set to 100 and 10, respectively. Similar to the first simulation, a total of 100 epoch was used for modeling training. We denoted our proposed prediction model as DNN-transfer, where the parameters associated with pre-trained DNNs and the newly added hidden layers were fixed and estimated, respectively. For comparison purposes, we also analyzed each simulated dataset using a DNN model with the same set of pre-selected genes and the same network architecture, except that all model parameters, including both the parameters in pre-trained models and those from the newly added hidden layers, are retrained. We denote this DNN model as DNN-optimal. We further analyzed each simulated dataset using existing widely adopted genomic risk prediction methods, including gBLUP [33] implemented in the gcta software, MultiBLUP [10], KMLMM [11] and DPR [15]. For each of these methods, we used their default setting (denoted as Default) as well as selected genes based on SKAT-linear, SKAT-optimal and ACAT.

Fig 4 (S2 Fig) and Fig 5 (S3 Fig) summarized the prediction accuracy under different screening thresholds for continuous and binary outcomes, respectively. As expected, when the outcomes are affected by genetic variants with non-linear effects (i.e., disease models *S*_2_ and *S*_3_), our proposed DNN-transfer significantly outperforms those prediction models that primarily focus on linear relationships (i.e., gBLUP, MultiBLUP and DPR). Although MKLMM is designed to capture non-linear effects through adopting a data-driven approach to select appropriate kernels, its performance can vary substantially depending on whether the appropriate kernels have been selected. For example, MKLMM can have similar level of performance as DNN-transfer when the most appropriate kernels have been selected. However, MKLMM can perform substantially worse once the selected kernels do not reflect the underlying relationships. When only linear additive effects are present (disease model *S*_1_), our DNN-transfer outperforms gBLUP under the default setting, but its performance is similar or slightly better than the other methods. This clearly indicates that similar to all deep learning models, our proposed DNN-transfer has natural advantages of capturing features with non-linear predictive effects, and it offers the flexibility in modeling features of high complexity. As shown in Fig 4 and Fig 5, the proposed DNN-transfer has very robust performance across a range of disease models, including a simple linear additive model to a more complex setting that involves different types of non-linear effects.

**Fig 4.**
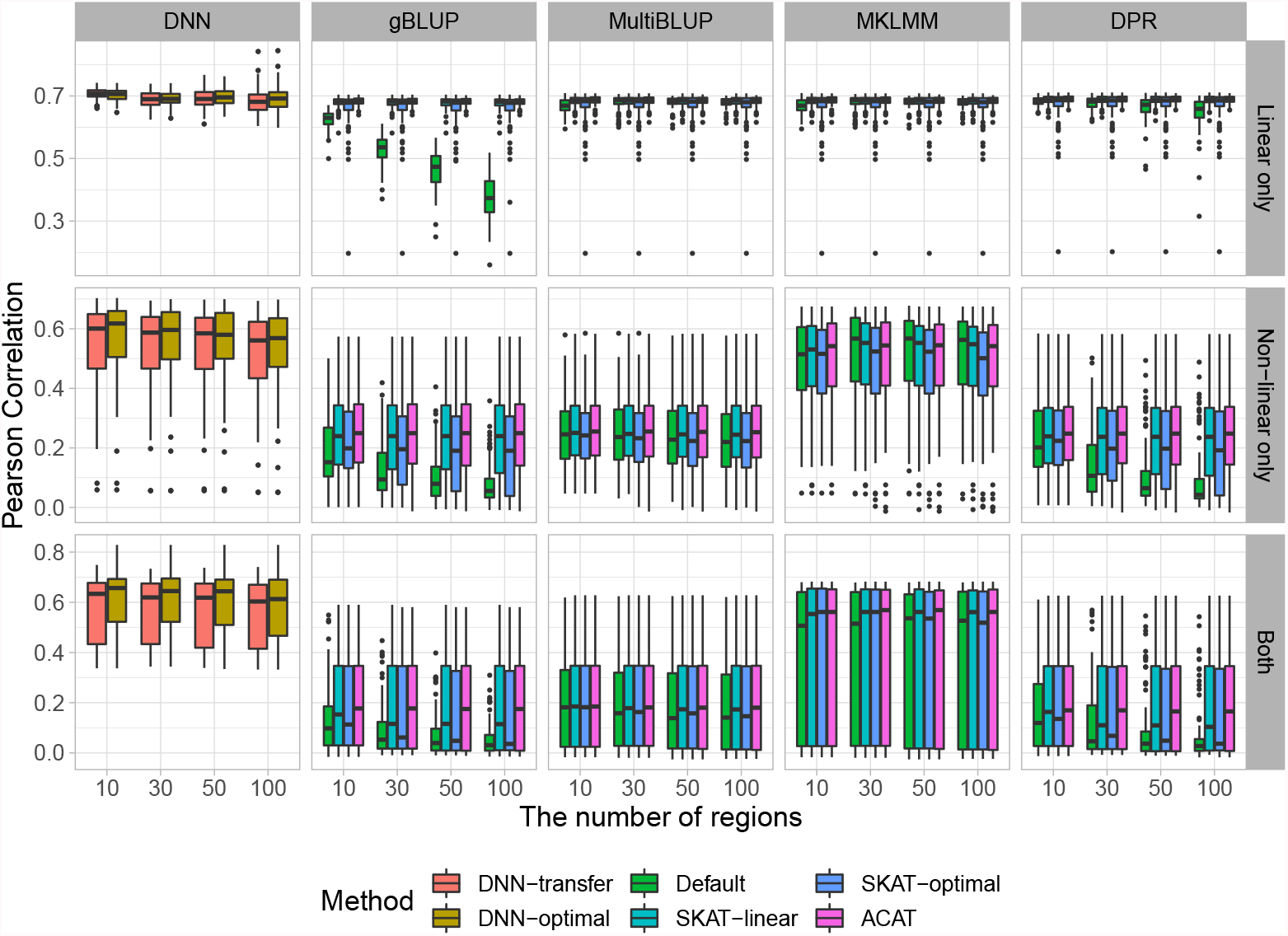
The comparisons of prediction accuracy for continuous outcomes. Genes with p-values less than 0.001 are considered significant.

**Fig 5.**
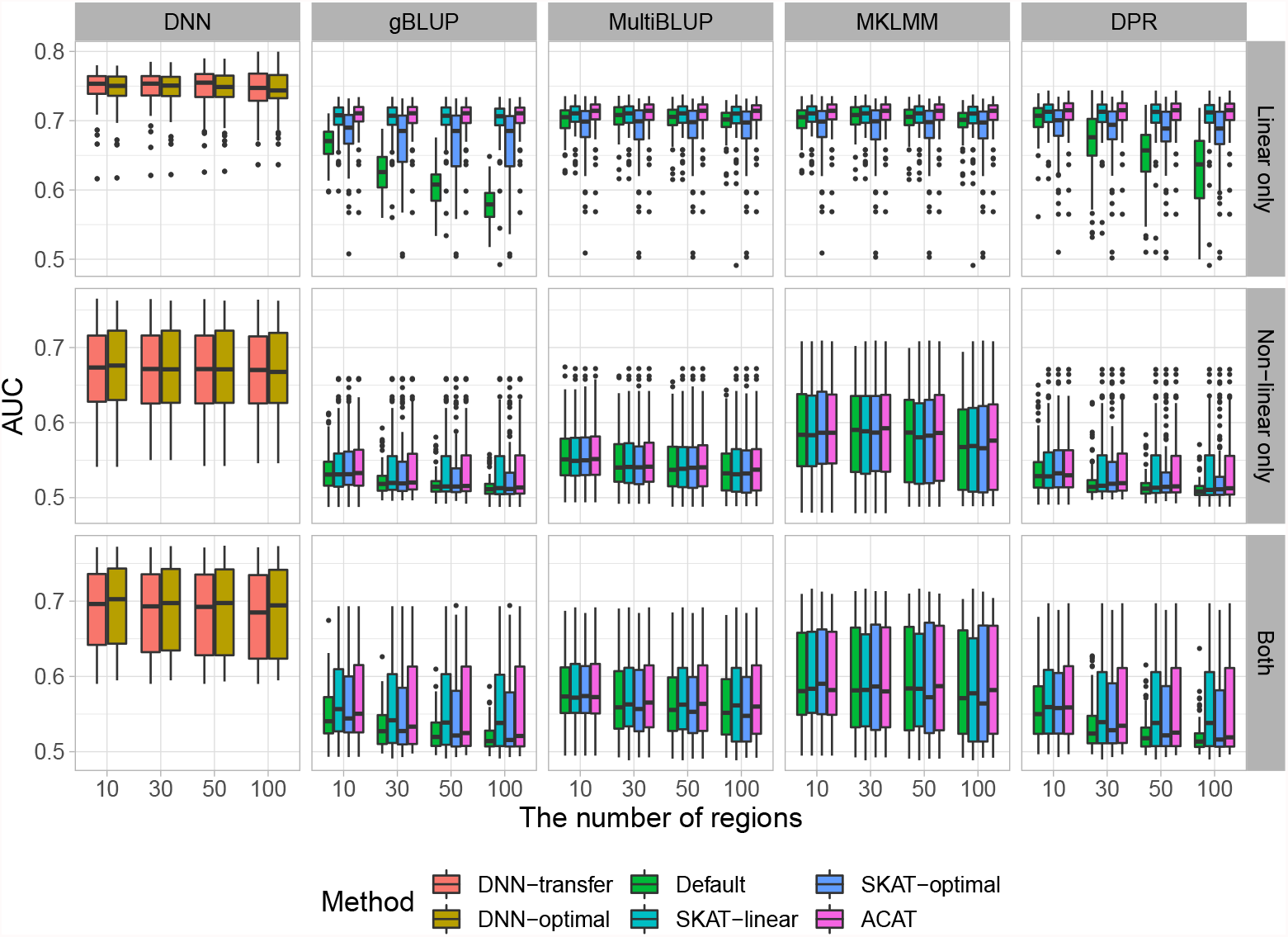
The comparisons of prediction accuracy for binary outcomes. Genes with p-values less than 0.001 are considered significant.

As the number of noise regions increases, the prediction accuracy for all methods that have employed a feature selection mechanism remains relatively stable, whereas the performance of those without feature selection (i.e., the default settings of gBLUP and DPR) dropped substantially. gBLUP assumes effect sizes from all genetic variants follow the same normal distribution, and the default setting of gBLUP include all genetic variants without filtering out the impact of noise. As shown in both Fig 4 and Fig 5, the prediction accuracy dropped the most for gBLUP under its default setting as the number of noise increases. Similarly, DPR sets its prior using a Dirichlet process with the stick-breaking constructive representation, and thus it models the effect sizes using a infinite normal mixture (i.e., 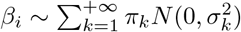). Although mixture models have the ability to model various types of effect size distributions, it cannot adequately tease out the impact of noise. Therefore, the prediction performance of DPR is affected by the amount of noise under its default setting. MultiBLUP, MKLMM, and our method employed their own feature screening processes by default, and thus as expected their performance remains largely unchanged as the number of noise increases. The robustness against noise can be critical for the prediction analysis, especially when analyzing high-dimensional genomic data.

For comparison purposes, in addition to the default settings, we also analyzed each simulated data using the above software (i.e., gBLUP, MultiBLUP, MKLMM, DPR), where predictive genetic regions are selected using commonly adopted methods, including SKAT-linear, SKAT-optimal and ACAT. As expected, with feature screening implemented, the performance for all methods remains stable as the number of noise increases. However, although SKAT-linear, SKAT-optimal and ACAT can efficiently identify features with linear effects and facilitate the downstream prediction tasks, their ability in detecting features with complicated types of effects is limited and thus the corresponding prediction models lack sufficient accuracy in the presence of non-linear effects. On contrary, our method first efficiently detects genes harboring both linear and non-linear effects, and then uses these selected genes to build DNN models. Our method can reduce the impact of noise via feature screening, and maximize prediction accuracy through modeling selected features with various types of effects. Therefore, it is robust against noise, regardless of the underlying disease models. We consider this important, as high-dimensional genomic data has a large amount of noise, and causal variants as well as their types of effects are unknown in advance.

Comparing the two DNN-based models (i.e., DNN-transfer v.s. DNN-optimal), although our proposed transfer learning method (i.e, DNN-transfer) has significantly reduced the number of model parameters, its prediction performance is very similar to DNN-optimal, where all parameters are re-estimated (Figs 4-5 and S2-S3 Figs). The proposed DNN-transfer utilizes the information from feature screening and only estimates the parameters associated with the newly added hidden layers (i.e., green box in Fig 1). Therefore, DNN-transfer substantially improves memory and computational efficiencies of a deep network that has the same architecture, making it scalable for the analysis of high-dimensional data. Indeed, in our proposed prediction framework, the small DNN models built for each gene during the feature screening process can be efficiently carried out via parallel computing, and the final prediction model directly uses these DNN models without re-training their associated parameters. Therefore, our proposed prediction framework has the capacity for the analysis of genome-wide data.

### Real data application

We applied the proposed prediction framework to analyze the whole-genome sequencing data obtained from ADNI, a multi-center longitudinal study aiming at detecting Alzheimer’s disease (AD) at the earliest stage and tracking AD progression with biomarkers [30]. Whole genome sequencing data from study participants in ADNI-2, including newly recruited and ADNI-1/GO continuing subjects, were obtained and analyzed using Illumina Genotyping Assays. After discarding genetically related individuals, 808 subjects remained in our study. Clinical, imaging and biospecimen biomarkers from these study participants were also collected. For our analyses, we focus on predicting the baseline PET imaging outcomes, including AV45 and FDG scans, using high-dimensional genomic data, where individuals with missing outcomes were excluded. The distributions of AV45 and FDG are shown in S4 Fig.

For the genome-wide data, we first filtered out genetic variants if they meet any of the following criteria: 1) call rate per subject ≤ 90%; 2) call rate per genetic variant ≤ 90%; 3) the p-value of Hardy-Weinberg equilibrium test ≤ 10^−5^; and 4) there is no variations for the genetic variant. After quality control, we annotated the remaining variants based on GRch37 assembly. A total of 21,985 genes with 18,087,684 genetic variants remained in our analyses, and the distribution of their minor allele frequencies is shown in S5 Fig.

### Feature selection for AV45 and FDG

We first used our proposed group-wise feature importance score to detect genes that are predictive for AV45 and FDG. For the network, it is constructed the same as our simulation, where a multi-layer perceptron with 2 hidden layers (*n*_1_ = 50 and *n*_2_ = 10) and a dropout layer is used. A total of 100 epoch was used for each model training. For comparison purposes, similar to simulation studies, we also used 1) SKAT with a linear kernel, 2) optimal SKAT and 3) the ACAT to detect phenotype-related features. The QQ-plots for AV45 and FDG are shown in S6 Fig and S7 Fig, respectively. Both the proposed DNN-screen and SKAT with a linear kernel controlled the type I errors well for both AV45 and FDG, whereas both SKAT-optimal and ACAT tend to have slightly inflated type I errors for the analyses of FDG. The Manhattan plots for the proposed DNN-screen and the other three methods are shown in Fig 6 and S8 Fig, respectively. While SKAT-optimal failed to identify any genes at the significance level of 10^−5^, all the other methods have identified three genes, including *APOE, APOC1* and *TOMM40*. All these three genes are located on chromosome 19, and they are well know AD-related genes [38–42]. For example, *APOE* encodes the Apolipoprotein E that plays an important role in the pathogenesis of AD. *APOE ϵ*_4_ is a major risk factor for AD in several populations (e.g., Caucasian and African American), and it is over-represented among late-onset AD patients [43, 44]. *APOC1* encodes the Apolipoprotein C1, and it involves in the cholesterol metabolism which can affect AD pathology. The *rs4420638* polymorphism on *APOC1* has an impact on the accumulation of homocysteine, which is involved in AD development and progression [45]. The *rs11568822* polymorphism on *APOC1* also increases the risk of AD in Caucasians, Asians, and Caribbeans [41]. *TOMM40* has been reported to be associated with late-onset AD, where the mitochondrial dysfunction is believed to be the underlying cause [39]. *rs10524523* on *TOMM40* affects the oxidative damage and thus influences the onset and progression of AD [45]. ACAT also detected *PVRL2* gene that is associated with both AV45 and FDG. However, since the type I errors for ACAT method tends to be inflated (S7 Fig), this additional gene should be interpreted with cautions.

**Fig 6.**
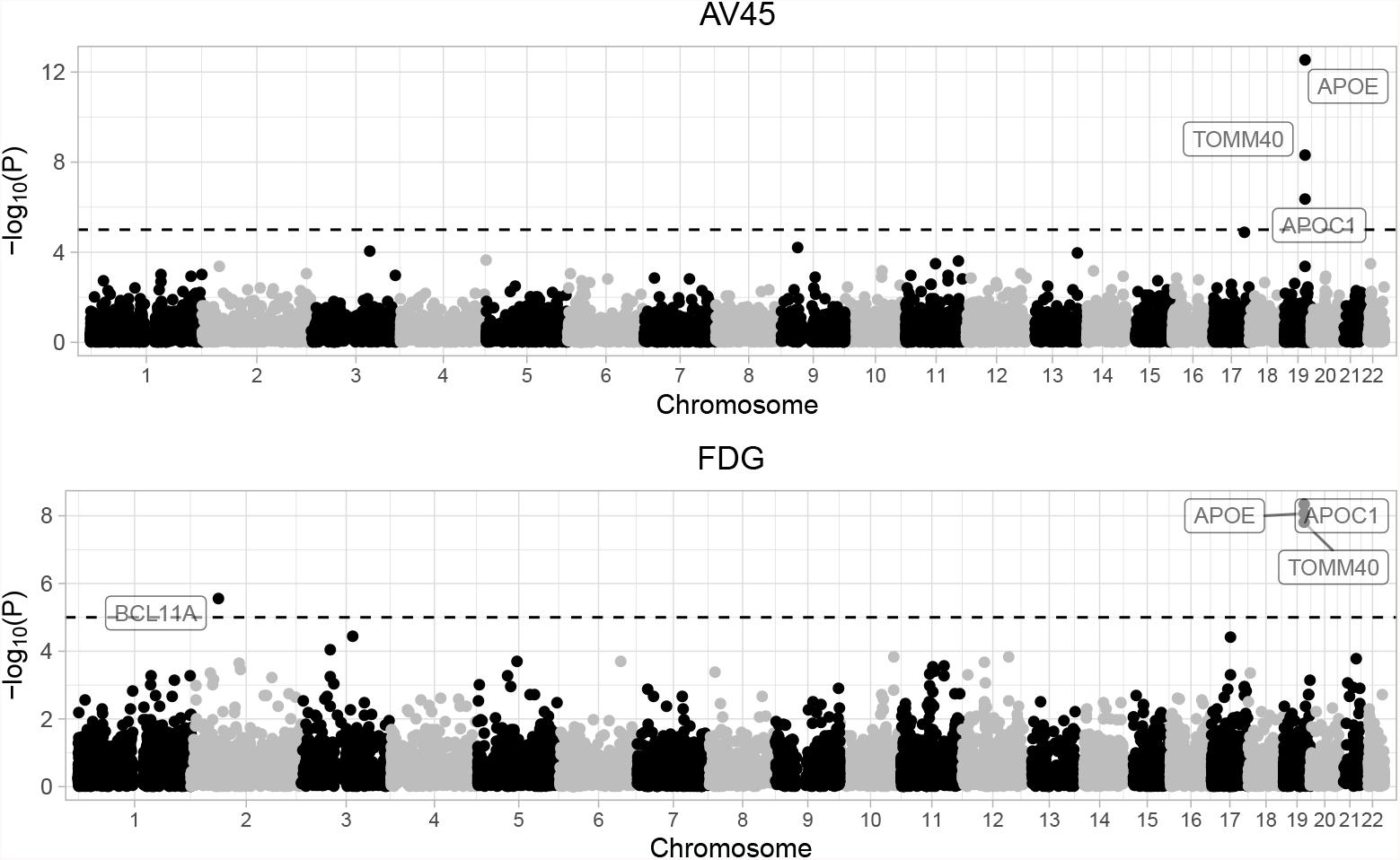
The Manhattan plot for AV45 and FDG using the DNN-screen method.

### The prediction analyses for AV45 and FDG

We further used our proposed DNN-framework to predict AV45 and FDG using the whole-genome data, where DNN-screen is employed to detect predictive genes and DNN-transfer is used to efficiently build prediction models with these selected genes. To reduce over-fitting, we randomly selected 100 individuals to serve as the testing set for each outcome, and used the remaining samples to select predictive genes and train prediction models. To avoid the chance finding, we repeated this process 20 times and reported the average prediction accuracy that is calculated based on the testing samples.

For the newly added hidden layers for our DNN-transfer model, it is set the same as our simulation studies, where 2 hidden layers with 100 and 10 hidden nodes for the first and second layers respectively is included. 100 epoch was used for each model training. For comparison purposes, we analyzed the same dataset using DNN-optimal, where the network architecture is the same as that in DNN-transfer and all parameters are re-estimated. We also analyzed each dataset using the widely-adopted gBLUP, MultiBLUP, MKLMM and DPR methods, where four scenarios were considered, including all genes as well as genes selected by SKAT-linear, SKAT-optimal and ACAT.

The pre-selected genes by DNN-screen, SKAT-linear, SKAT-optimal and ACAT for each sample are summarized in S3 Table. Using *p <* 10^−5^ as the threshold, DNN-screen, SKAT-linear and ACAT methods selected *APOE, APOC1* and *TOMM40* as predictive genes, and the chances for selecting them are all above 85%. All the other genes except *PVRL2* are selected no more than 5%. The SKAT-optimal did not detect any genes at this threshold level. For *PVRL2* gene, the chances of being selected as predictive genes by DNN-screen and ACAT are 30% and 100%, respectively. However, the chance of detecting *PVRL2* as predictive for SKAT-based methods is 0. The genes that are selected as predictive genes under other thresholds are listed in S3 Table.

The prediction accuracy when genes are pre-selected under the thresholds of 0.001 and 0.005 is shown in Fig 7 and S9 Fig, respectively. For both AV45 and FDG, the DNN-based models (i.e., DNN-transfer and DNN-optimal) have better prediction accuracy than all the other methods considered, including their default settings and those with feature screening implemented. For the prediction of AV45, under the threshold of 0.001, gBLUP, MultiBLUP and MKLMM perform similarly, whereas the accuracy of DPR tends to be slightly worse. For the prediction of FDG, MultiBLUP, MKLMM and DPR perform similarly, whereas gBLUP tends to have better performance than them. Among feature screening methods compared, ACAT usually provides the best accuracy for each of the prediction model considered (i.e., gBLUP, MultiBLUP, MKLMM and DPR). While pre-selection can reduce the impact of noise, they can also overlook the impact of SNPs with infinitesimal effects. That can be part of the reasons why ACAT tends to perform better than SKAT-based methods. As shown in S6 Fig and S7 Fig, ACAT usually selects more features than the others, making it easier to capture SNPs with small effects. However, given ACAT could have inflated type I error, the models built with features selected by the ACAT can be hard to interpret. For our proposed DNN-based models, we have accommodated two widely used model assumptions in the network architecture design, where the background input node is designed to model the infinitesimal effects and the selected gene nodes are used to capture the sparse predictive effects. Therefore, as shown in Fig 7 and S9 Fig, the proposed DNN architecture has better prediction accuracy.

**Fig 7.**
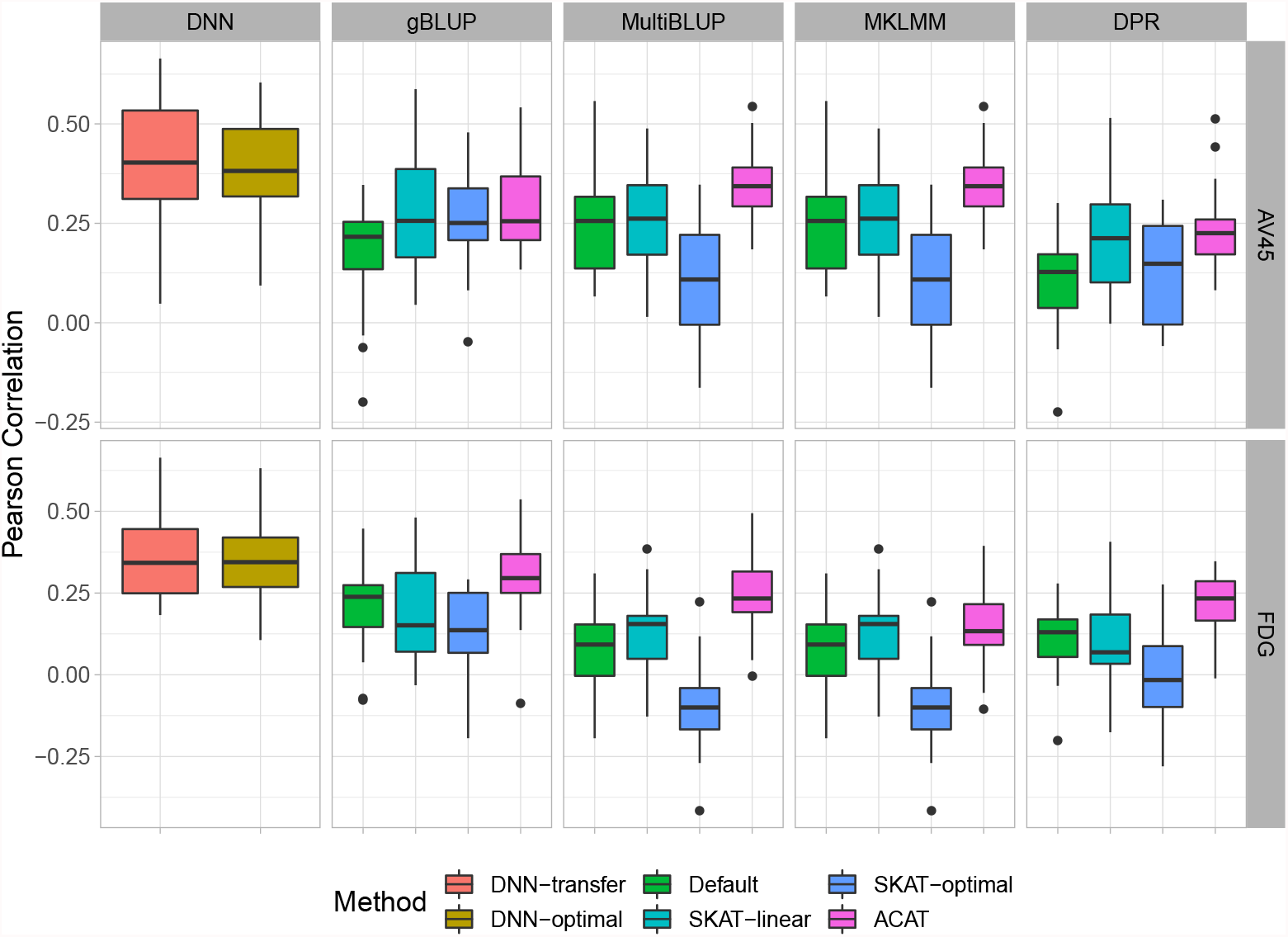
The prediction accuracy for AV45 and FDG. Genes are pre-selected under the p-value threshold of 0.001 for DNN-transfer, SKAT-linear, SKAT-optimal and ACAT.

Comparing the two DNN-based models, while our proposed DNN-transfer uses the idea of transfer learning and has significantly reduced the model complexity, its prediction performance is very similar to the DNN model that re-trains all model parameters. Therefore, although the proposed DNN-transfer is much more computationally efficient as compared to a DNN model with the same architecture, it can still maintain a similar level of accuracy.

## Discussion

In this paper, we proposed a novel deep neural network based prediction framework for the analysis of high-dimensional genomic data. We first proposed an explainable group-wise feature importance score, which can efficiently detect predictive genes with various types of effects and control the type I error well. We then developed a computationally efficient deep transfer learning model for prediction analysis, where information from feature screening is directly incorporated and both linear and non-linear predictive effects can be efficiently captured. Through extensive simulation studies and real data analyses, we have shown that our proposed framework is more powerful in detecting predictive genes and it has better prediction accuracy, especially when the predictive effects are non-linear.

Dimension reduction is critical for analyzing genome-wide data. Existing widely-used methods usually treat dimension reduction and prediction modeling in two separate steps with different objective functions [9, 13]. Therefore, they can overlook important features, leading to a sub-optimal prediction model. Recently developed methods tend to build prediction models with the feature selection embedded [12, 20]. However, these methods can barely be applied to genome-wide data due to their high computational cost. Furthermore, existing feature selection methods usually focus on linear relationships, and thus is unlikely to identify predictors with non-linear effects. Our proposed modeling framework overcomes the above limitations by streamlining the feature selection and prediction modeling processes, where both aim at capturing predictive effects of various forms and maximizing the prediction accuracy (i.e., the objective functions are the same). In particular, our screening process selects genes that are “significantly” predictive. It first builds a DNN model for each gene, where the prediction accuracy is maximized. It then detects predictive genes that harbor both linear and non-linear effects through comparing the prediction accuracy from DNN models built with a set of features and their permutations. Our proposed DNN-screen aligns well with the downstream prediction task, and is unlikely to overlook features that are highly predictive. We used a DNN model for feature screening, which is mainly because our prediction model is also DNN-based. Indeed, other machine learning models (e.g., support vector machine and random forest) can also be employed for the feature screening process, where the group-wise feature importance score is calculated by comparing the prediction accuracy of the model with original features and their permutations. In addition, we have derived the distribution of our proposed group-wise feature importance score based on the data splitting idea, and the computationally expensive procedure that requires to refit the model for each permuted data is not needed, making it possible to consider models of high complexity (e.g., deep neural network with complex architecture).

Substantial amount of evidences have suggested that non-linear predictive effects widely exist [16]. However, existing literature for the calculation of PRS usually focuses on linear relationships [6, 10, 13, 15, 20], ignoring the contributions from predictors with non-linear effects (e.g., interaction effects). Kernel functions have been recently incorporated into the prediction model to capture those non-linear effects [11, 17], but their performance highly depends on the pre-selected kernels and the underlying disease model. In our proposed modeling framework, we developed a transfer-learning-based deep neural network for the PRS construction. Therefore, our prediction model inherits all the advantages in deep neural networks, and can discover and model features of high complexity. Different from deep neural network that usually suffers from the curse of dimensionality and high computational cost, our proposed DNN-transfer uses the idea of transfer learning and directly incorporates information from feature screening into the network architecture. Therefore, it significantly reduced the number of model parameters as compared to models of a similar level of complexity, making it much more appealing in handling high-dimensional data. Indeed, our proposed framework can be applied to genome-wide data, where feature screening is used to scan all genetic variants and a prediction model is built by directly incorporating information from feature screening into the final prediction task.

While the proposed DNN-transfer utilizes the idea from transfer learning [46], it has its own unique characteristics. Traditional transfer learning method utilizes a pre-trained model that is obtained by jointly considering all features. However, for high-dimensional data, these pre-trained models themselves can be hard to obtain, mainly due to their huge amount of model parameters and the high computational cost. On contrary, our proposed method builds the pre-trained model (i.e., blue box of Fig 1) by combining multiple gene-based DNN models, which are trained separately in the feature screening process. The average computational time for feature screening as sample sizes increases is illustrated in S10 Fig. As the gene-based DNN models built in the feature screening process can also be efficiently implemented using parallel computing, the pre-trained model in our proposed framework can be easily obtained. The final architecture of our proposed prediction model includes a pre-trained model module that is obtained by combining multiple pre-trained DNNs (Fig 1: blue box), and an added hidden layer module that is designed to capture joint predictive effects from these genes (Fig 1: green box). Similar to transfer learnings, we keep the parameters in the pre-trained model fixed and only estimate the parameters associated with newly added hidden layers, which has substantially reduced the model complexity while maintaining their capability in capturing predictive effects of various forms. Indeed, for a genome-wide analysis with approximately 22,000 genes and a sample size of 500 (i.e., the analysis of FDG and AV45), our feature screening takes about 15 seconds for each gene and the transfer-learning based prediction model takes no more than 5 minutes for the final prediction tasks. Through both simulations (Figs 4-5, S2-S3 Fig) and real data analyses (Fig 7 and S9 Fig), we have shown that our proposed DNN-transfer can obtain similar levels of prediction accuracy as compared to a similar DNN with all parameters re-estimated. Therefore, DNN-transfer can jointly consider a large number of genes and efficiently build an accurate prediction model.

Model interpretation can be of great importance in the field of Bioinformatics. While deep learning models have achieved the state-of-the-art prediction performance in many domains, their black-box nature limited their applications for risk prediction studies. Unlike many existing DNNs (e.g., autoencoder and convolutional neural network), our proposed DNN-based feature screening and prediction modeling framework has much better interpretability. Our proposed group-wise feature importance score can detect predictive genes that is a functional unit of DNA, and our designed prediction network architecture can reflect the underlying disease etiology. While we used the proposed group-wise feature importance score to detect predictive genes, the same idea can also be used to detect disease-associated pathways. In addition, although we mainly focus on the prediction analysis based on genomic data in this work, our method can be applied to the analysis of various other data types (e.g., multi-omics data), where the proposed DNN-based prediction framework is first used to detect the complex inter/intra-relationships among multi-omcis data within a set (e.g., pathway) and then build prediction models by using the detected predictive sets.

In the prediction analyses of FDG and AV45 using whole-genome data, we have found that our proposed framework has achieved better prediction accuracy than existing methods (Fig 7 and S9 Fig). It consistently selected *APOE, APOC1* and *TOMM40* as highly predictive genes, and all of them are well-known AD related. For example, it has been shown that genetic polymorphisms in *APOE* and *APOC1* genes are associated with cognitive impairment progression in patients with late-onset AD [41, 43, 44, 47]. Evidences have also suggested that *APOE ϵ*_4_ itself increases cognitive decline, and *APOC1 H2* has a synergistic effect with *APOE ϵ*_4_ in increasing the risk of cognitive decline [47]. *rs2075650* polymorphism on *TOMM40* gene contributes to AD in Caucasian and Asian populations [48]. The polymorphic poly-T variant *rs10524523* on *TOMM40* gene provides better estimation of age of late-onset AD for *APOE ϵ*_3_ carriers [49]. While our genome-wide analyses improve the prediction accuracy and offer more insight, additional replication studies are needed to further investigate these risk prediction models and their utilities.

One of the limitations of our proposed prediction framework is that we carried out feature screening at the gene level. Therefore, similar to many existing methods that only consider marginal effects and within-gene interaction effects [11], our method can overlook genes with only between gene interaction effects. A potential solution to this problem is that feature screening can be implemented at the pathway levels, where interactions between genes within the pathway can be explicitly modeled. To eliminate the impact of genes that are not predictive within the pathway, a variational dropout layer can be added into our proposed transfer learning-based prediction model [50]. This can be a future direction of our research.

In summary, we have developed a DNN-based prediction modeling framework, which can not only discover predictive features of complex forms, but also accurately and efficiently build an explainable prediction model that can capture features of high complexity. The proposed modeling framework is among the first few DNN-based method that can be applied to genome-wide data, and it is implemented in a python package that can be obtained from https://github.com/YaluWen/EDNN.

## Data Availability

All data produced in the present study are available upon reasonable request to the authors

## Supporting information

The supporting information can be found in supplementary.pdf.

## Acknowledgments

We wish to acknowledge the contribution of NeSI high-performance computing facilities to the results of this research. This project is funded by the National Natural Science Foundation of China (Award No. 82173632 and 81903418), Early Career Research Excellence Award from the University of Auckland, and the Marsden Fund from Royal Society of New Zealand (Project No. 19-UOA-209).

