## Supplementary Materials for "Explainable deep transfer learning model for disease risk prediction using high-dimensional genomic data"

#### Supplementary Tables

**S1 Table. Effect sizes for four causal genes in simulation 1**

| Outcomes | Sample Size | $\beta_1$ | $\beta_2$ | $\beta_3$ | $\beta_4$ |
|---|---|---|---|---|---|
| **Continuous** | $N = 1000$ | $N(0, 2^2)$ | $N(0, 8^2)$ | 0.5 | 10 |
| | $N = 10000$ | $N(0, (1/3)^2)$ | $N(0, (4/3)^2)$ | 0.3 | 10 |
| **Binary** | $N = 1000$ | $N(0, 2^2)$ | $N(0, 8^2)$ | 0.5 | 10 |
| | $N = 10000$ | $N(0, 2^2)$ | $N(0, 8^2)$ | 0.5 | 10 |

**S2 Table. Effect sizes for four causal genes in simulation 2**

| Disease model | $\beta_1$ | $\beta_2$ | $\beta_3$ | $\beta_4$ |
|---|---|---|---|---|
| $S_1$: **Linear effects only** | $N(0, 0.5^2)$ | $N(0, 2^2)$ | 0 | 0 |
| $S_2$: **Non-linear effects only** | 0 | 0 | 0.125 | 10 |
| $S_3$: **Both linear and non-linear effects** | $N(0, 0.5^2)$ | $N(0, 1)$ | 0.125 | 5 |

**S3 Table.** The probability of genes being selected (only genes that have been selected more than 50% under the significance level of 0.001 are listed)

| Gene | Chromosome | $P < 0.005$ | $P < 0.001$ | $P < 10^{-5}$ |
|---|---|---|---|---|
| | | *DNN-screen* | | |
| APOC1 | 19 | 0.950 | 0.900 | 0.850 |
| APOE | 19 | 1.000 | 1.000 | 1.000 |
| PVRL2 | 19 | 0.900 | 0.750 | 0.300 |
| TOMM40 | 19 | 0.950 | 0.850 | 0.850 |
| | | *SKAT-linear* | | |
| SPRR2G | 1 | 1.000 | 0.700 | 0.000 |
| TGFB2 | 1 | 1.000 | 0.550 | 0.000 |
| LINC00471 | 2 | 1.000 | 0.700 | 0.000 |
| DLD | 7 | 1.000 | 0.600 | 0.000 |

Continued on next page

**S3 Table.** The probability of genes being selected (only genes that have been selected more than 50% under the significance level of 0.001 are listed)

| **Gene** | **Chromosome** | $P < 0.005$ | $P < 0.001$ | $P < 10^{-5}$ |
|---|---|---|---|---|
| ADAM28 | 8 | 1.000 | 1.000 | 0.000 |
| CPB2-AS1 | 13 | 1.000 | 0.950 | 0.000 |
| CPB2 | 13 | 1.000 | 0.950 | 0.000 |
| LINC01070 | 13 | 1.000 | 1.000 | 0.000 |
| FBXO33 | 14 | 0.950 | 0.800 | 0.000 |
| RPA1 | 17 | 1.000 | 0.700 | 0.000 |
| APOC1 | 19 | 1.000 | 1.000 | 1.000 |
| APOE | 19 | 1.000 | 1.000 | 1.000 |
| NTF4 | 19 | 1.000 | 0.900 | 0.050 |
| TOMM40 | 19 | 1.000 | 1.000 | 0.950 |
| LOC101928269 | 21 | 1.000 | 0.800 | 0.000 |
| | *SKAT-optimal* | | | |
| ATR | 3 | 1.000 | 0.800 | 0.000 |
| APOC3 | 11 | 1.000 | 0.950 | 0.000 |
| PAFAH1B2 | 11 | 1.000 | 0.650 | 0.000 |
| SIK3 | 11 | 1.000 | 0.900 | 0.000 |
| MEDAG | 13 | 1.000 | 0.600 | 0.000 |
| BCAR1 | 16 | 1.000 | 0.900 | 0.000 |
| KRTAP22-1 | 21 | 0.900 | 0.550 | 0.000 |
| LSS | 21 | 1.000 | 1.000 | 0.000 |
| | *ACAT* | | | |
| ANKRD20A12P | 1 | 0.800 | 0.700 | 0.000 |
| URB2 | 1 | 0.850 | 0.750 | 0.000 |
| ZBTB8B | 1 | 0.850 | 0.850 | 0.000 |
| ZBTB8OS | 1 | 0.850 | 0.550 | 0.000 |
| SCHLAP1 | 2 | 0.850 | 0.750 | 0.000 |
| CWH43 | 4 | 0.800 | 0.650 | 0.000 |
| PPP1R2P3 | 5 | 1.000 | 0.850 | 0.000 |
| HIST1H3F | 6 | 0.900 | 0.900 | 0.000 |
| FAM133B | 7 | 0.800 | 0.600 | 0.000 |
| FAM133DP | 7 | 0.800 | 0.600 | 0.000 |
| OMD | 9 | 0.750 | 0.550 | 0.000 |
| CADM1 | 11 | 0.800 | 0.750 | 0.000 |
| LOC101054525 | 11 | 0.800 | 0.800 | 0.000 |
| RNF214 | 11 | 0.900 | 0.550 | 0.000 |
| CAND1 | 12 | 0.900 | 0.700 | 0.000 |
| F10 | 13 | 0.800 | 0.800 | 0.000 |
| LINC01070 | 13 | 1.000 | 0.900 | 0.000 |
| GALK2 | 15 | 0.800 | 0.600 | 0.000 |
| TMED3 | 15 | 0.850 | 0.850 | 0.000 |
| AFMID | 17 | 0.800 | 0.800 | 0.000 |
| ENDOV | 17 | 0.900 | 0.550 | 0.000 |
| SHMT1 | 17 | 0.850 | 0.850 | 0.000 |

Continued on next page

**S3 Table.** The probability of genes being selected (only genes that have been selected more than 50% under the significance level of 0.001 are listed)

| **Gene** | **Chromosome** | $P < 0.005$ | $P < 0.001$ | $P < 10^{-5}$ |
|---|---|---|---|---|
| TK1 | 17 | 0.800 | 0.800 | 0.000 |
| APOC1 | 19 | 1.000 | 1.000 | 1.000 |
| APOE | 19 | 1.000 | 1.000 | 1.000 |
| PVRL2 | 19 | 1.000 | 1.000 | 1.000 |
| TOMM40 | 19 | 1.000 | 1.000 | 1.000 |
| ZNF146 | 19 | 0.700 | 0.550 | 0.000 |
| ZNF565 | 19 | 0.950 | 0.900 | 0.050 |
| LOC101928269 | 21 | 1.000 | 0.750 | 0.050 |

### Supplementary Figures

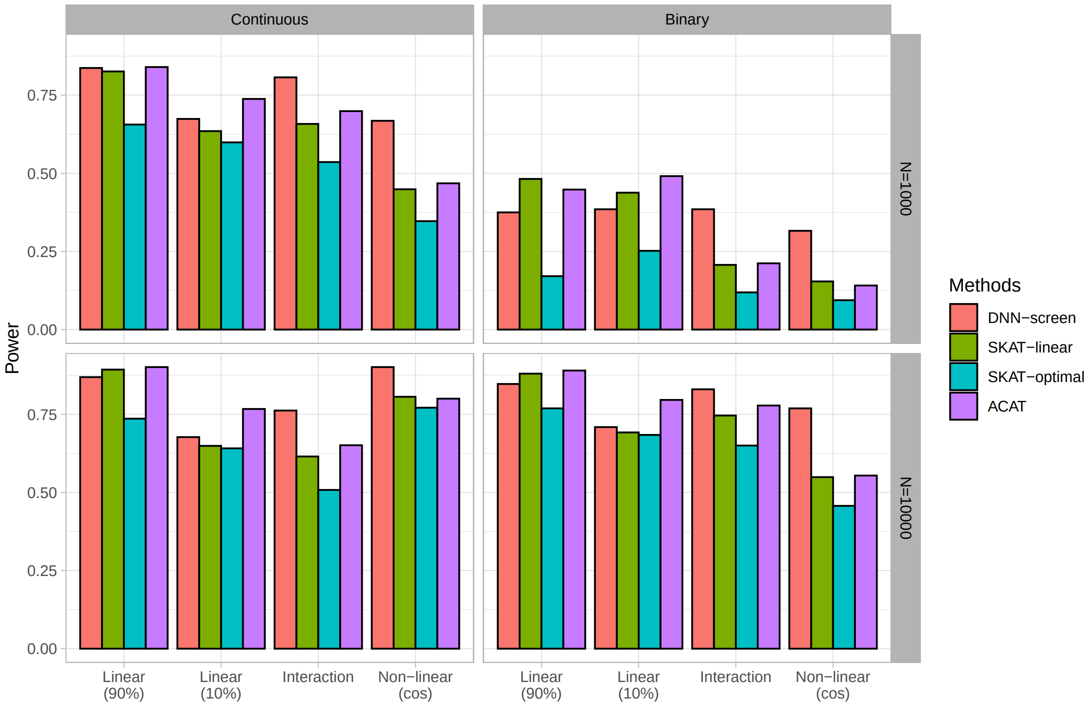


**S1 Fig. The comparisons of power under 1% significance level.** Linear (90%): 90% of genetic variants on the causal gene is predictive. Linear (10%): 10% of genetic variants on the causal gene is predictive. Interaction: pairwise interaction effects. Non-linear (cos): genetic variants on the causal gene affect the outcome through a cosine function.

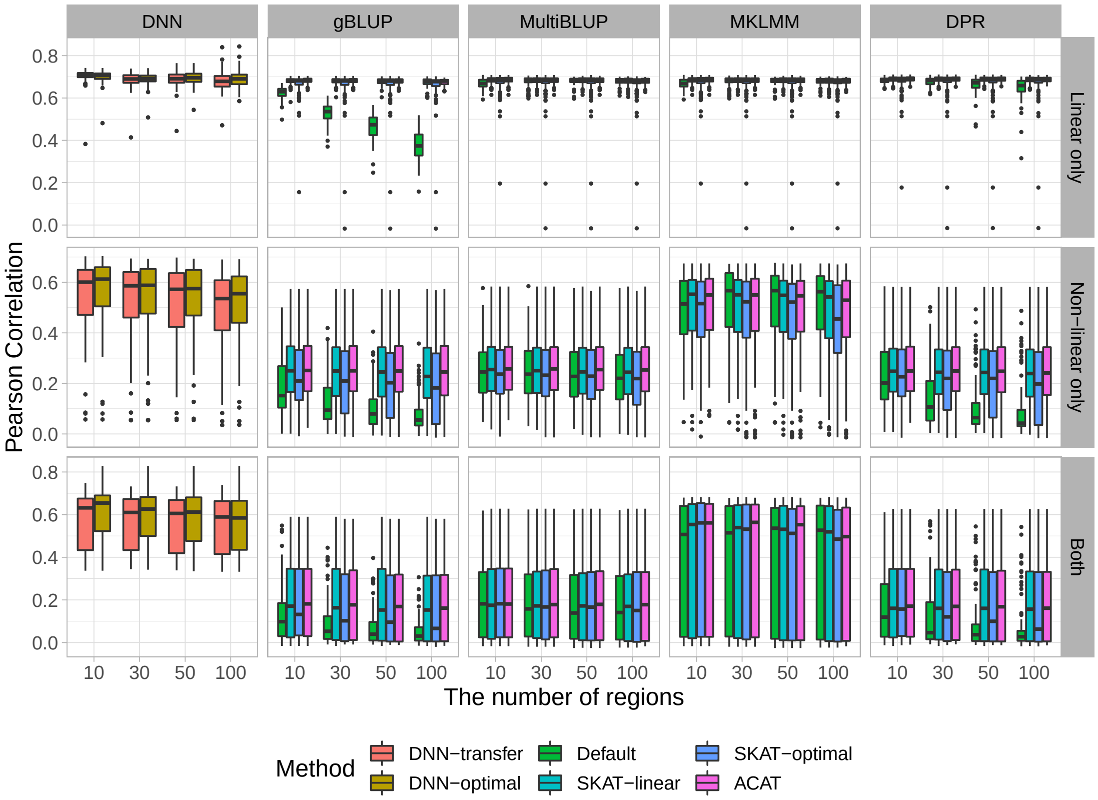


**S2 Fig.** The comparisons of prediction accuracy for continuous outcomes. Genes with p-values less than 0.005 are considered significant.

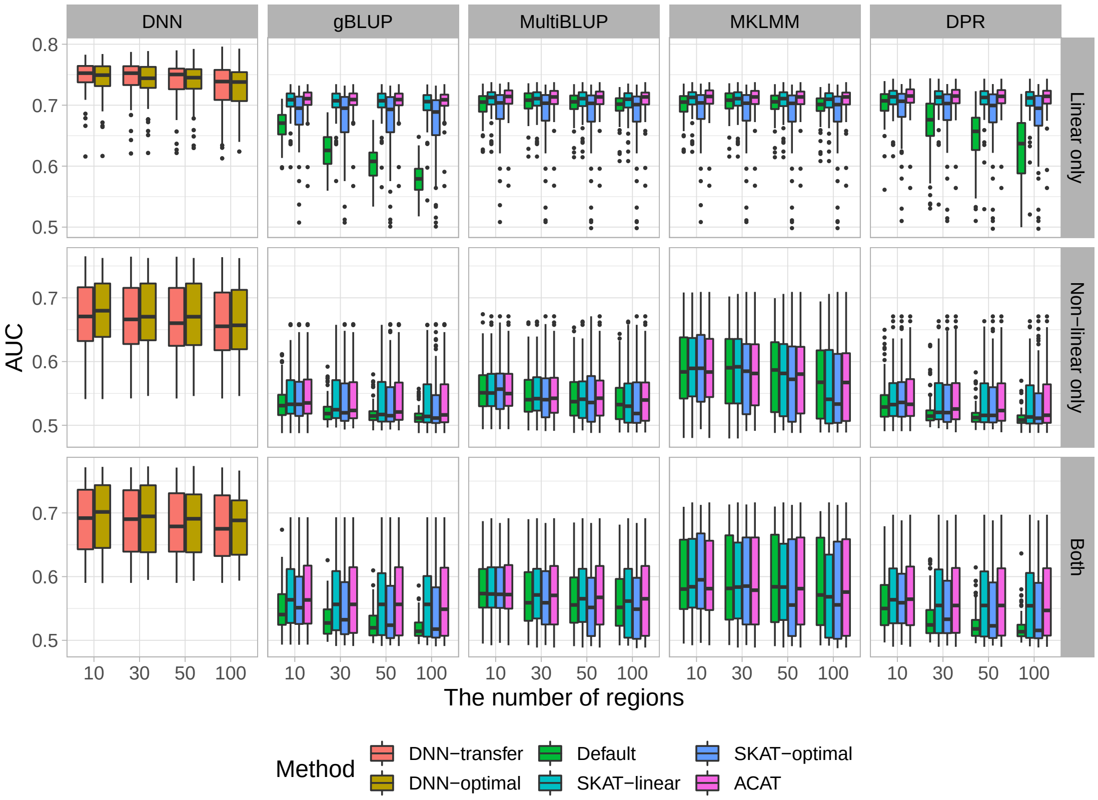


**S3 Fig.** The comparisons of prediction accuracy for binary outcomes. Genes with p-values less than 0.005 are considered significant.

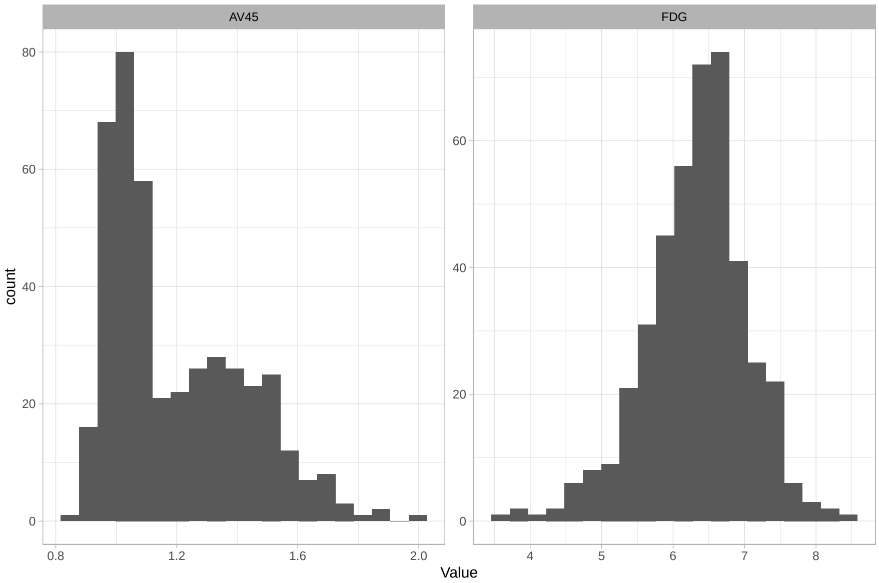


**S4 Fig.** The distributions of AV45 and FDG.

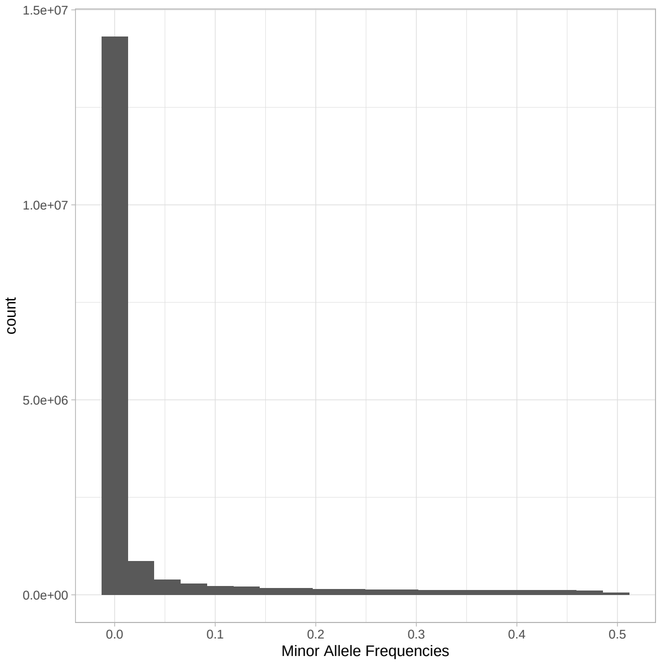


**S5 Fig.** The distribution of minor allele frequencies

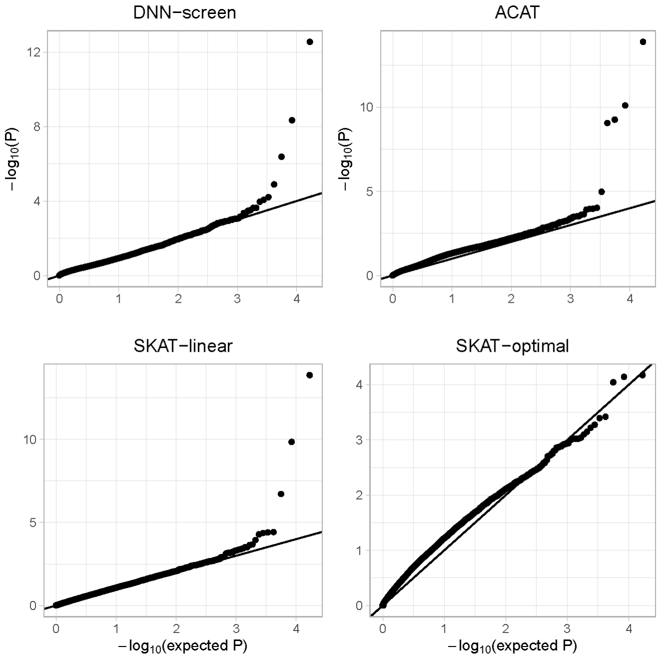


**S6 Fig.** QQ-plot for AV45

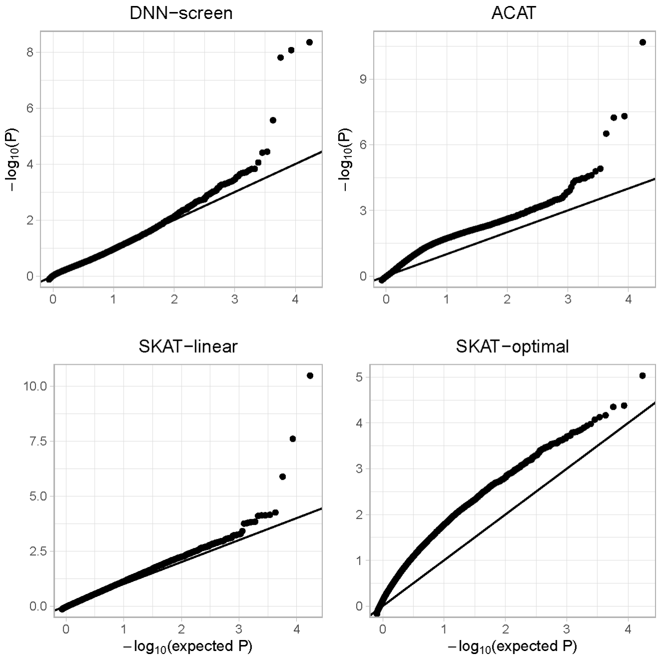


**S7 Fig.** QQ-plot for FDG

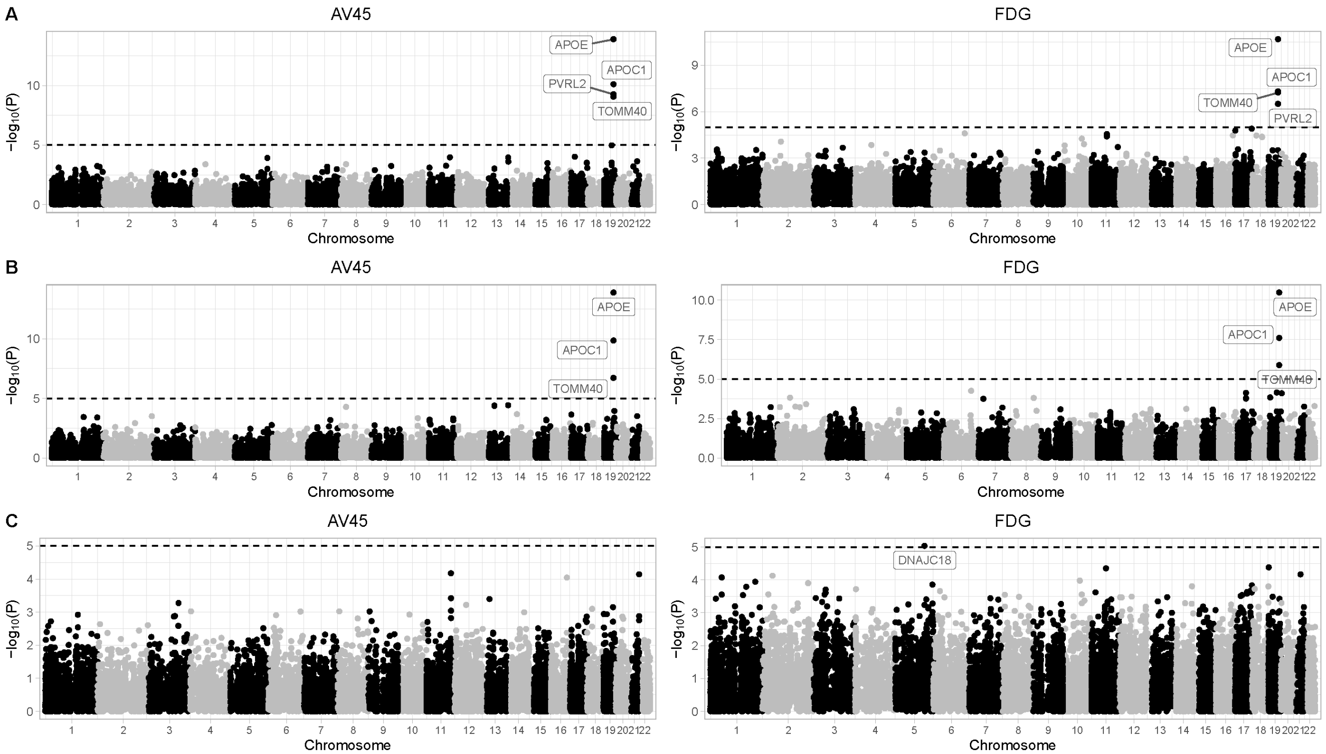


**S8 Fig.** The Manhattan plot for AV45 and FDG using the ACAT, SKAT-linear and SKAT-optimal methods. **A**: ACAT method. **B**: SKAT with linear kernel. **C**: SKAT hat optimally combines the burden test and SKAT.

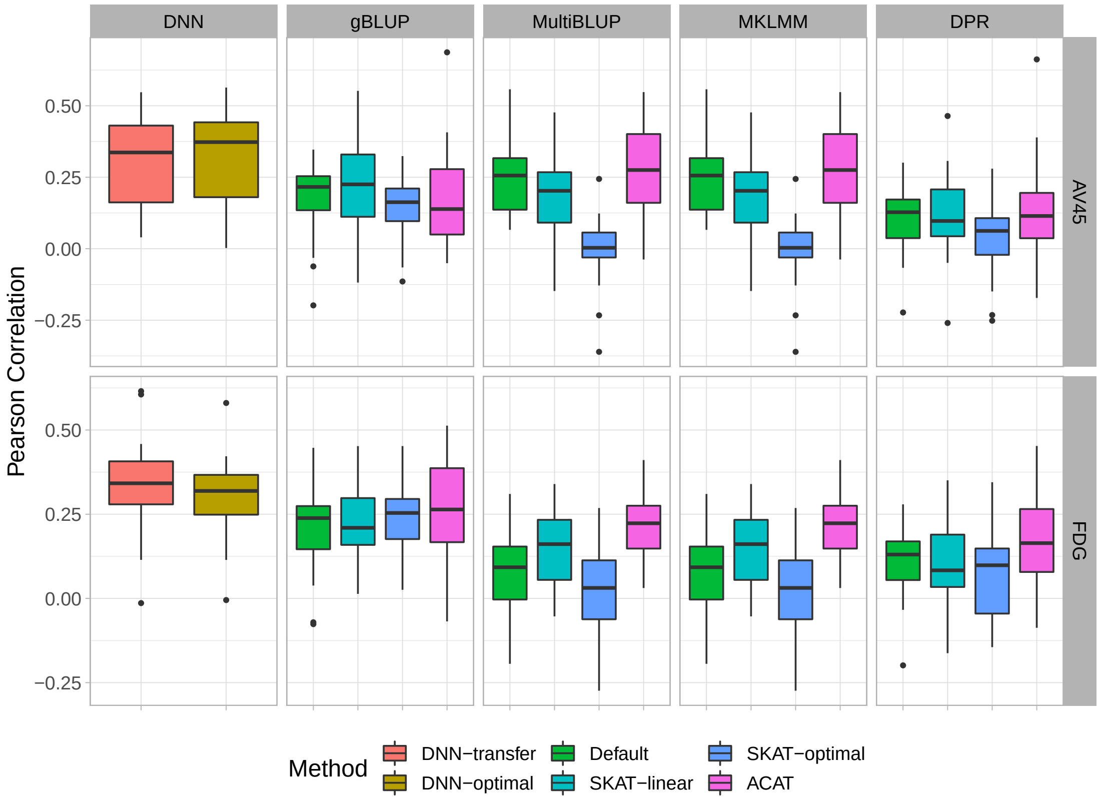


**S9 Fig.** The prediction accuracy for AV45 and FDG. Genes are pre-selected under the p-value threshold of 0.005 for DNN-transfer, SKAT-linear, SKAT-optimal and ACAT

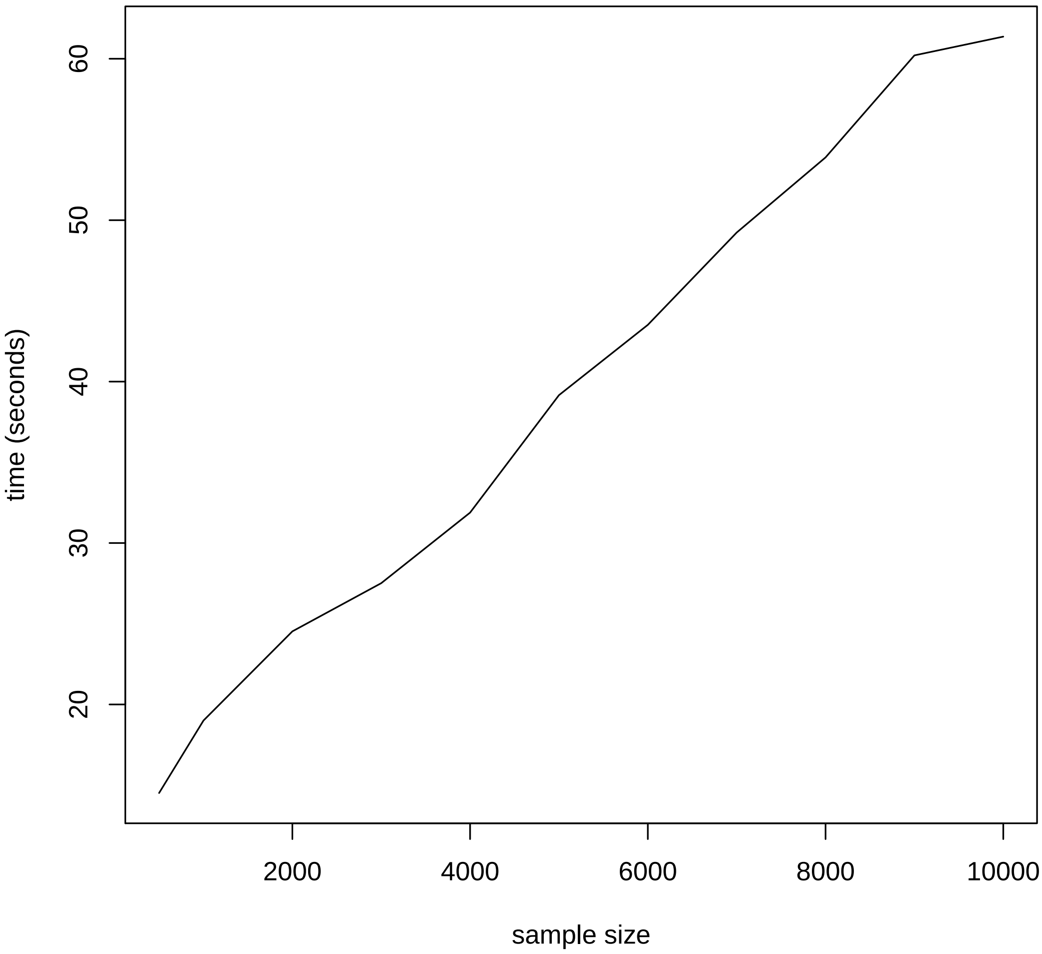


**S10 Fig.** The average computational time for feature screening as sample sizes increase
